# Durability of immune responses to the booster mRNA vaccination against COVID-19

**DOI:** 10.1101/2022.12.02.22282921

**Authors:** Prabhu S. Arunachalam, Lilin Lai, Hady Samaha, Yupeng Feng, Mengyun Hu, Harold Sai-yin Hui, Bushra Wali, Madison Ellis, Christopher Huerta, Kareem Bechnack, Sarah Bechnack, Matthew Lee, Matthew Litvack, Cecilia Losada, Alba Grifoni, Alessandro Sette, Veronika I. Zarnitsyna, Nadine Rouphael, Mehul S. Suthar, Bali Pulendran

## Abstract

Waning immunity to vaccination represents a major challenge in vaccinology. Whether booster vaccination improves the durability of immune responses is unknown. Here we show, using a cohort of 55 adult vaccinees who received the BNT162b2 (Pfizer-BioNTech) or mRNA-1273 (Moderna) vaccine against SARS-CoV-2, that a booster (i.e., 3^rd^ immunization) dose at 6 - 10 months increased the half-life of serum neutralizing antibody (nAb) titers to 76 days from 56 - 66 days estimated after the primary two-dose vaccination series. A second booster dose (i.e., 4^th^ immunization) more than a year after the primary vaccination increased the half-life further to 88 days. However, despite this modestly improved durability in nAb responses against the Wuhan strain, there was a loss in neutralization capacity against Omicron subvariants, especially the recently emerged variants, BA.2.75.2 and BQ.1.1 (35 and 50-fold drop in titers respectively, relative to the ancestral (WA.1) strain. While only 55 – 65% of participants demonstrated a detectable nAb titer against the newer variants after the booster (3^rd^ dose), the response declined to below the detection limit in almost all individuals by 6 months. Notably, even against BA.1 and BA.5, the titers declined rapidly in a third of the vaccinees and were below the detection limit at 6 months. In contrast, booster vaccination induced antigen-specific memory B and T cells that persisted for at least 6 months. Collectively, our data show that the durability of immune responses improves following subsequent booster immunizations; however, the emergence of immune evasive variants reduces the effectiveness of booster doses in preventing infection.

## INTRODUCTION

Recent studies have shown declining efficacy of the Pfizer-BioNTech and Moderna mRNA vaccines after primary and booster vaccinations (Andrews et al., 2022; Collie et al., 2022). Although the booster vaccination (3^rd^ dose) was effective in protection against severe disease (Lustig et al., 2022; Moreira et al., 2022), vaccine effectiveness against symptomatic disease declined rapidly to ^50% in the real world as a result of the emergence of the Omicron subvariants BA.4/BA.5 (Collie *et al*., 2022). The fourth dose showed no significant increase in efficacy against infection as compared to the third dose suggesting that further booster immunizations may only have a marginal benefit (Regev-Yochay et al., 2022). The underlying immunological basis for the declining efficacy is unknown.

We and others have demonstrated a rapid decline in serum antibody titers following a two-dose primary vaccination with BNT162b2 (Suthar et al., 2022) or mRNA1273 (Doria-Rose et al., 2021; Pegu et al., 2021) with a considerable number of individuals demonstrating weak or no nAb response against immune evasive viral variants. The half-life of serum nAb response was estimated to be 56 - 66 days up to 6 months after two doses of BNT162b2 or mRNA1273 (Doria-Rose *et al*., 2021; Goel et al., 2021; Suthar *et al*., 2022). Recent data indicate that the decline in antibody response may be slower after the 3^rd^ dose as compared to the 2^nd^ dose (Ailsworth et al., 2022); however, breakthrough infections significantly influence the antibody kinetics (Gilboa et al., 2022; Qu et al., 2022b). Here, we systematically evaluated the magnitude and durability of binding and nAb responses, and memory T and B cell responses after the 3^rd^ and 4^th^ doses of mRNA vaccination to determine the immunological mechanisms of declining vaccine efficacy.

## RESULTS

### Study design and participants

We recruited 55 volunteers who received a booster vaccination of BNT162b2 or mRNA1273 6 – 10 months post-completion of the primary series. Of the 55 individuals, 20 received mRNA1273 and 35 received BNT162b2. Additionally, we recruited 13 individuals who received their 4^th^ dose 12 – 20 months post the second dose and 6 – 8 months post the 3^rd^ dose. Of these 13 individuals, all but one individual received mRNA1273, and all are immunocompetent. A schematic of the study design is shown in **Fig. 1A**. The age, gender, race, vaccination, and breakthrough infections details of the participants are presented in **Supplementary Table 1**. In addition to the clinical report of infections, we measured anti-nucleocapsid (anti-N) antibody responses in all individuals to determine potential undiagnosed infection with SARS-CoV-2 **(Supplementary Fig. 1A and B)**. Thirty-one of the 55 volunteers who received the 3^rd^ dose and 7 of the 13 who received the 4^th^ dose were determined to be SARS-CoV-2 naïve. Of those individuals in the 3-dose cohort who had COVID-19, 10 participants had an anti-NC response prior to the booster, and 14 had a response after the booster. In the 4-dose cohort, 3 participants each had COVID-19 before and after the final booster.

**Figure 1.**
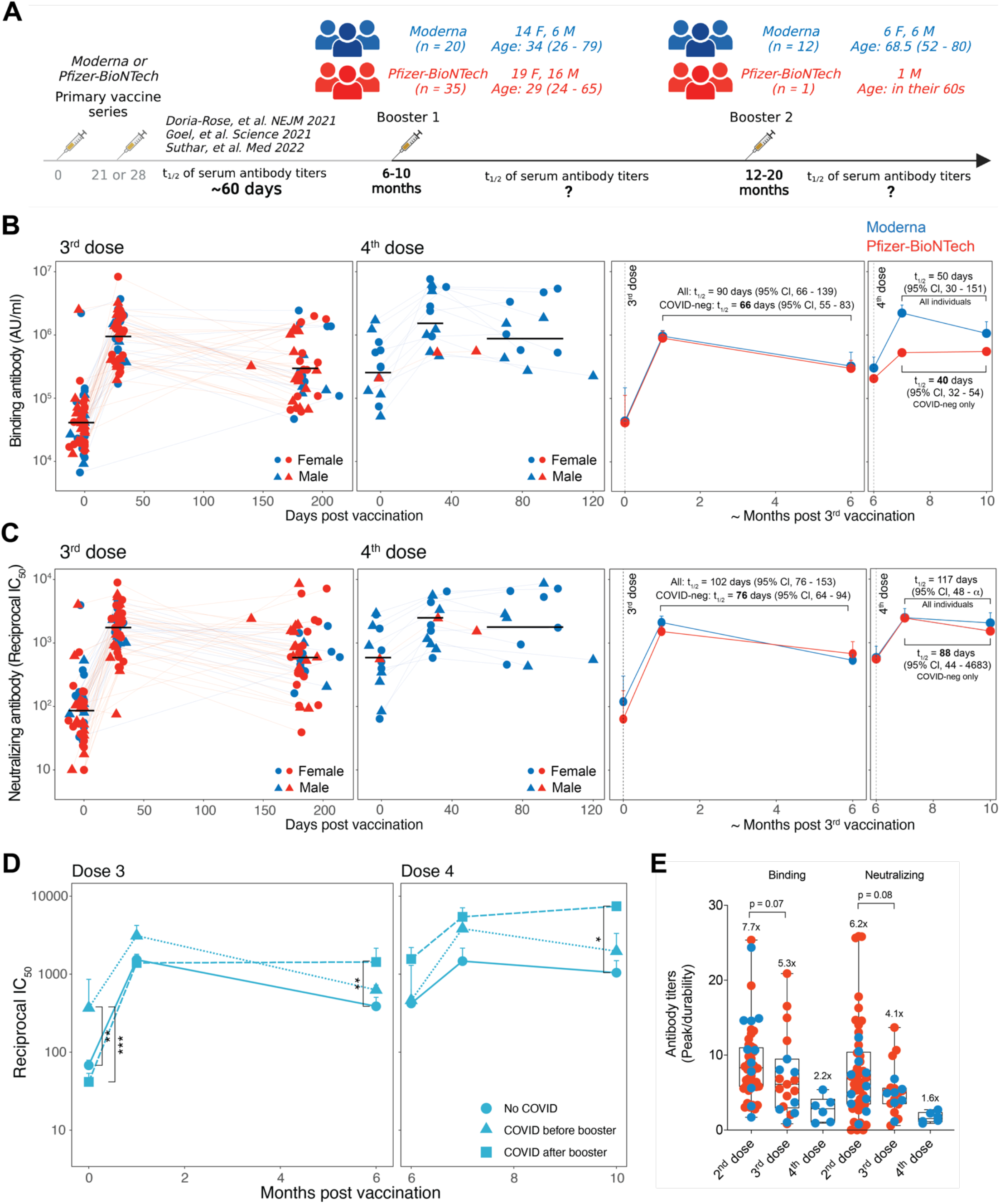
Serum antibody responses following booster mRNA vaccinations. **(A)** A schematic of the study design and participant details. (**B – C)** Anti-Spike binding IgG **(B)** and live-virus nAb titers **(C)** against the ancestral WA.1 strain. Each symbol represents an individual in the left two plots. The black horizontal lines indicate geometric mean titers. The two graphs on the right show summary (Geometric mean + SEM) of antibody responses. **(D)** Neutralizing antibody titers in groups of participants stratified by exposure to COVID-19. The data shown are geometric mean in each group + SEM. The statistical difference between groups at each time point was analyzed by the Mann-Whitney test. **(E)** Fold change between binding and neutralizing antibody responses measured at peak (^1 month) versus durability (^6 months) after 2^nd^, 3^rd^ or 4^th^ doses. Each symbol represents a SARS-CoV-2 naïve participant. COVID-19^+^ individuals were removed from the analysis. The statistical significance was determined using the Mann-Whitney test. The data of BNT162b2 after 2^nd^ vaccination were published previously by us.

### Durability of antibody responses to booster vaccinations

Booster vaccination (3^rd^ dose) elicited anti-Spike binding IgG titers in all individuals **(Fig. 1B, left panel)**. The geometric mean titers (GMT) increased 21-fold from 0.43 × 10^5^ AU/ml at the baseline to 9.18 × 10^5^ AU/ml in the first month. There was a 3-fold drop in the titers at 6 months when the GMT was 3 × 10^5^ AU/ml. The GMT at baseline of the individuals who received their 4^th^ dose (approximately 6 months after their 3^rd^ dose (Fig 1A)) was 2.9 × 10^5^ AU/ml, almost the same as that of the 6-month time point in the 3-dose group. The titers increased 6.7-fold (GMT: 19.9 × 10^5^ AU/ml) one month after the booster and decreased 2-fold (GMT: 10 × 10^5^ AU/ml) during the 3 – 4-month follow-up period **(Fig. 1B, middle left panel)**. The responses between mRNA1273 and BNT162b2 showed no significant difference **(Fig. 1B, panels on the right)**. The half-life of binding antibody titers estimated using the exponential decay model was 90 days when all individuals were considered **(Fig. 1B, middle right panel)**. However, the half-life estimate decreased to 66 days, when only those individuals defined as SARS-CoV-2 naïve were considered, in line with the evidence that the durability of humoral immune responses is influenced by breakthrough infection (Gilboa *et al*., 2022). The half-life estimates after the 4^th^ dose were 50 days or 40 days when considering all or SARS-CoV-2-naive individuals, respectively (**Fig. 1B, right panel**).

The live-virus nAb response was detectable against the ancestral strain at the time of the 3^rd^ dose in the majority of individuals with a GMT IC_50_ of 81. The titers increased 21-fold (GMT: 1713) and were sustained at a magnitude 7-fold higher than the pre-booster titers (GMT: 594) **(Fig. 1C, left panel)**. The nAb titers at the peak and 6-month time points were 5- and 12-fold higher, respectively, than titers at the corresponding time points after 2 doses measured in our previous studies using the same assay (Arunachalam et al., 2021; Suthar *et al*., 2022). The 4^th^ dose increased the GMT to 2477, which persisted without considerable decay (GMT: 1997) during the follow-up period **(Fig. 1C, middle left panel)**. The half-life calculated using the exponential decay model with all the individuals was 102 days, substantially higher than the estimate of 56 days up to 6 months after the primary mRNA vaccine series (Suthar *et al*., 2022). The half-life of the nAb response decreased to 76 days when only SARS-CoV-2 naïve individuals were analyzed, which was still significantly higher than the half-life of 56 days after the second dose (p = 0.003, Wald test). The half-life estimates after the 4^th^ dose were 117 days or 88 days when considering all or SARS-CoV-2-naive individuals, respectively (**Fig. 1C, right panel**). Consistent with these estimates, the nAb response in individuals who had a breakthrough infection after the booster vaccination persisted significantly higher than in SARS-CoV-2 naïve individuals **(Fig. 1D)**. Neither the magnitude nor the durability of responses was significantly different between males and females **(Supplementary Fig. 1C)**.

To compare the durability of antibody responses after 2^nd^, 3^rd^, and 4^th^ doses directly, we calculated the fold-change between peak versus durability time points of binding as well as nAb responses after each vaccination only in SARS-CoV-2 naïve individuals. The data after 2 vaccinations for BNT162b2 was previously published in our study (Suthar *et al*., 2022). The ratio improved marginally upon subsequent booster immunizations consistent with our half-life estimates **(Fig. 1E)**. Whether the antibody durability after the 4^th^ dose continues to persist until 6 months remains to be investigated. Notably, there was an inverse correlation between age and the peak versus durability fold-change, but not the peak magnitude suggesting a relatively higher persistence in older adults **(Supplementary Fig. 1D, E)**. Taken together, these data show that subsequent booster immunizations improve the half-life of antibody titers modestly, which in conjunction with the increase in absolute magnitude of nAb titers results in significantly improved durability of antibody responses.

### Antibody breadth

We and others have reported generation of antibodies that can neutralize Omicron following a booster mRNA vaccination (3^rd^ dose) (Edara et al., 2022; Garcia-Beltran et al., 2022; Pajon et al., 2022; Sievers et al., 2022). Consistent with these studies, we observed live-virus nAb titers against Omicron BA.1, BA.5, and BA.2.75 subvariants (**Fig. 2A)**. The peak GMTs were 133, 115, and 98 against BA.1, BA.5 and BA.2.75, respectively, 12 to 18-fold lower than that of the ancestral strain **(Fig. 2A)**. In contrast, the GMTs against the recently emerged variants of interest BA.2.75.2 and BQ.1.1 measured in a subset of 18 individuals with the highest nAb titers against other viruses were 39 and 24, respectively, with 30% – 45% of individuals below the limit of detection **(Fig. 2A, B)**. These titers were ^5-fold lower than against Omicron BA.1 or BA.5, and 35 to 50-fold lower than the response against WA.1 strain **(Fig. 2B)**. By 6 months, almost no one had a detectable response against BA.2.75.2 and BQ.1.1. Notably, even against BA.1 and BA.5, approximately a third of the vaccinees had no detectable nAb response by 6 months **(Fig. 2B, right panel)**. This observation prompted us to investigate whether a lack of exposure to SARS-CoV-2 is associated with a more rapid decline in nAb response against variants. Our analysis showed that the nAb titers against the variants at baseline were significantly higher in individuals who had COVID-19 prior to the booster vaccination **(Fig. 2C)**. One month after the booster, all individuals elicited nAbs against the variants, but the response persisted durably (p < 0.05) in individuals who had a breakthrough infection after the booster **(Fig. 2C)**. It was notable that the nAb titers in individuals who had COVID-19 prior to the booster declined to levels statistically comparable to that of SARS-CoV-2 naïve vaccinees despite eliciting relatively higher titers following the booster. We further investigated the durability of nAbs in only SARS-CoV-2 naive participants and observed that those individuals who rapidly declined to below the detection limit against the variants also had lower titers against the ancestral strain at all time points **(Fig. 2D)**. Although the magnitude of nAb titers was only ^2-fold lower in this subgroup, the decline in response was rapid resulting in weak, and in the case of the variants, no neutralization titers at 6 months. Finally, we also measured nAb titers against BA.1, BA.5, and BA.2.75 after the 4^th^ dose **(Fig. 2E)**. Consistent with nAb titers against WA.1 **(Fig. 1C)**, variant-specific nAb titers were relatively higher at baseline but induced more moderately by vaccination (4^th^ dose) and persisted stably until the follow-up period at 3 to 4 months **(Fig. 2E)**.

**Figure 2.**
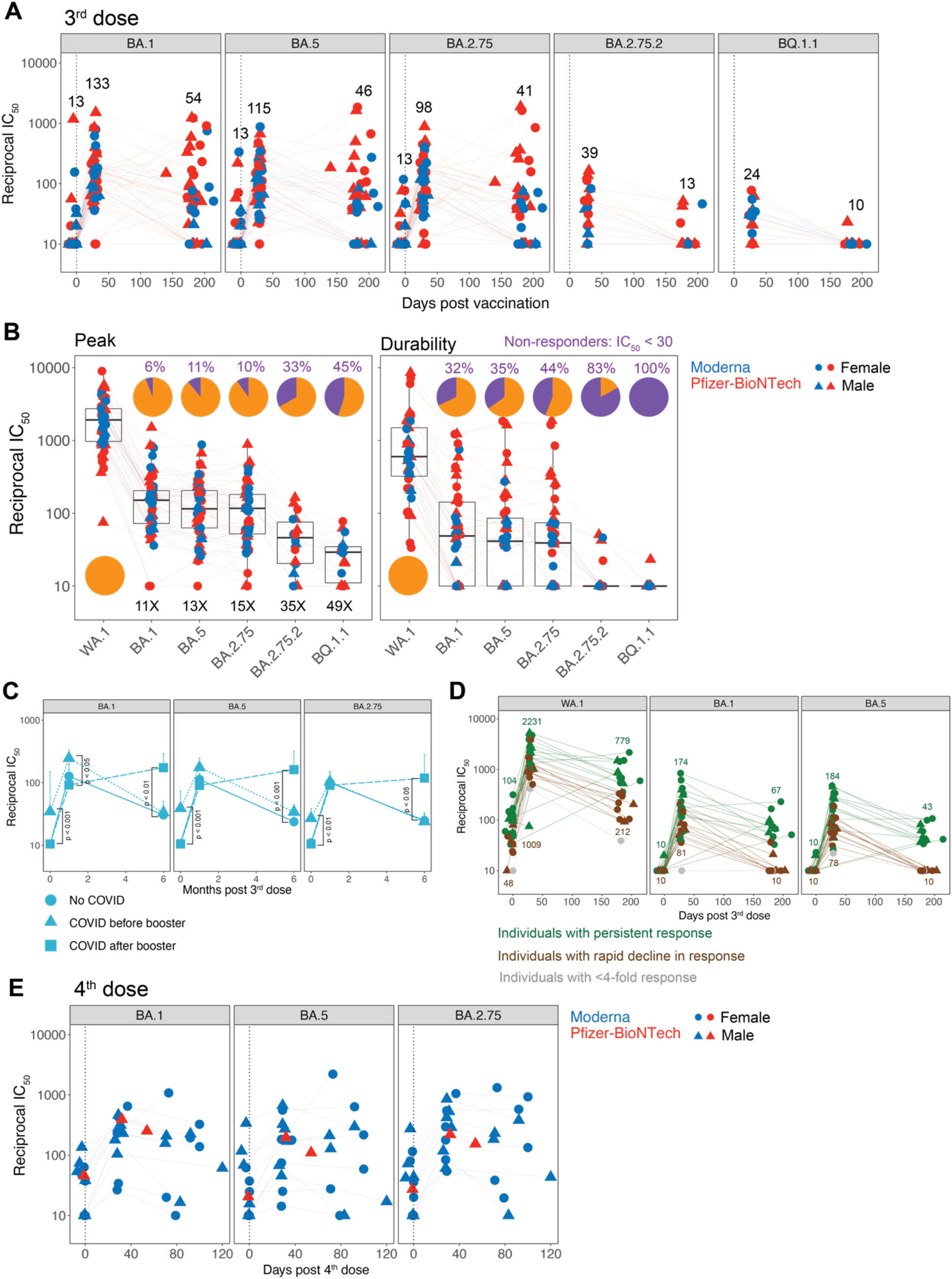
Neutralizing antibody breadth following booster vaccinations. **(A)** Live-virus neutralizing antibody response measured against Omicron BA.1, BA.5, BA.2.75, BA.2.75.2 and BQ.1.1. (N = 55 for BA.1, BA.5 and BA.2.75; N = 18 for BA.2.75.2 and BQ.1.1). Participants who had the highest titers against the ancestral and Omicron BA.1 variants were selected for this assay. **(B)** Neutralizing antibody titers against the viruses indicated on X-axis at peak (left panel) and durability (right panel). The pie charts show the proportion of participants who responded against each virus. Non-responders were defined as those who had an IC_50_ below 30. The numbers inside the graph followed by X show the decrease in titers against variants in comparison to the ancestral strain. **(C)** Neutralizing antibody titers against the variants indicated on the plots in participants stratified by exposure to COVID-19. The data shown are geometric mean in each group + SEM. The statistical difference between groups at each time point was analyzed by the Mann-Whitney test. **(D)** Neutralizing antibody titers in all SARS-CoV-2 naïve individuals. Each symbol represents an individual. Individuals who showed a neutralization titer < 20 against BA.1, BA.5, or BA.2.75. at 6 months were classified as those with rapid decline (brown). Individuals who showed a < 4-fold increase in titers against the variants were colored in grey. The rest of the individuals were considered normal responders (green). **(E)** Live-virus neutralizing antibody response measured against Omicron BA.1, BA.5, BA.2.75, BA.2.75.2 and BQ.1.1. (N = 13) in participants who received the 4^th^ dose of the mRNA vaccine.

### Cellular immune responses

Next, we assessed Spike-specific memory B cells by flow cytometry analysis of PBMCs labeled with ?uorescently-tagged recombinant Spike and RBD (receptor-binding domain) proteins (**Fig. 3A**). The 3^rd^ dose vaccination significantly increased the frequency of Spike-binding memory B cells from 1.9% (of CD20^+^ IgM^-^ IgD^-^ IgA^-^ IgG^+^) at baseline to 3.2% at ^1 month. The response persisted durably and was maintained at 2.6% six months later **(Fig. 3B, top left panel)**. The frequency of RBD^+^ memory B cells followed the same kinetics but was present at a lower magnitude in comparison to Spike-binding B cells **(Fig. 3B, bottom panel)**. While we did not have the appropriate peak time point to observe a potential increase in the magnitude after the 4^th^ dose, the memory B cell frequency persisted at the pre-vaccination magnitude by 3-4 months **(Fig. 3B)**. The memory B cell frequencies were not significantly associated with sex **(Supplementary Fig. 2)**.

**Figure 3.**
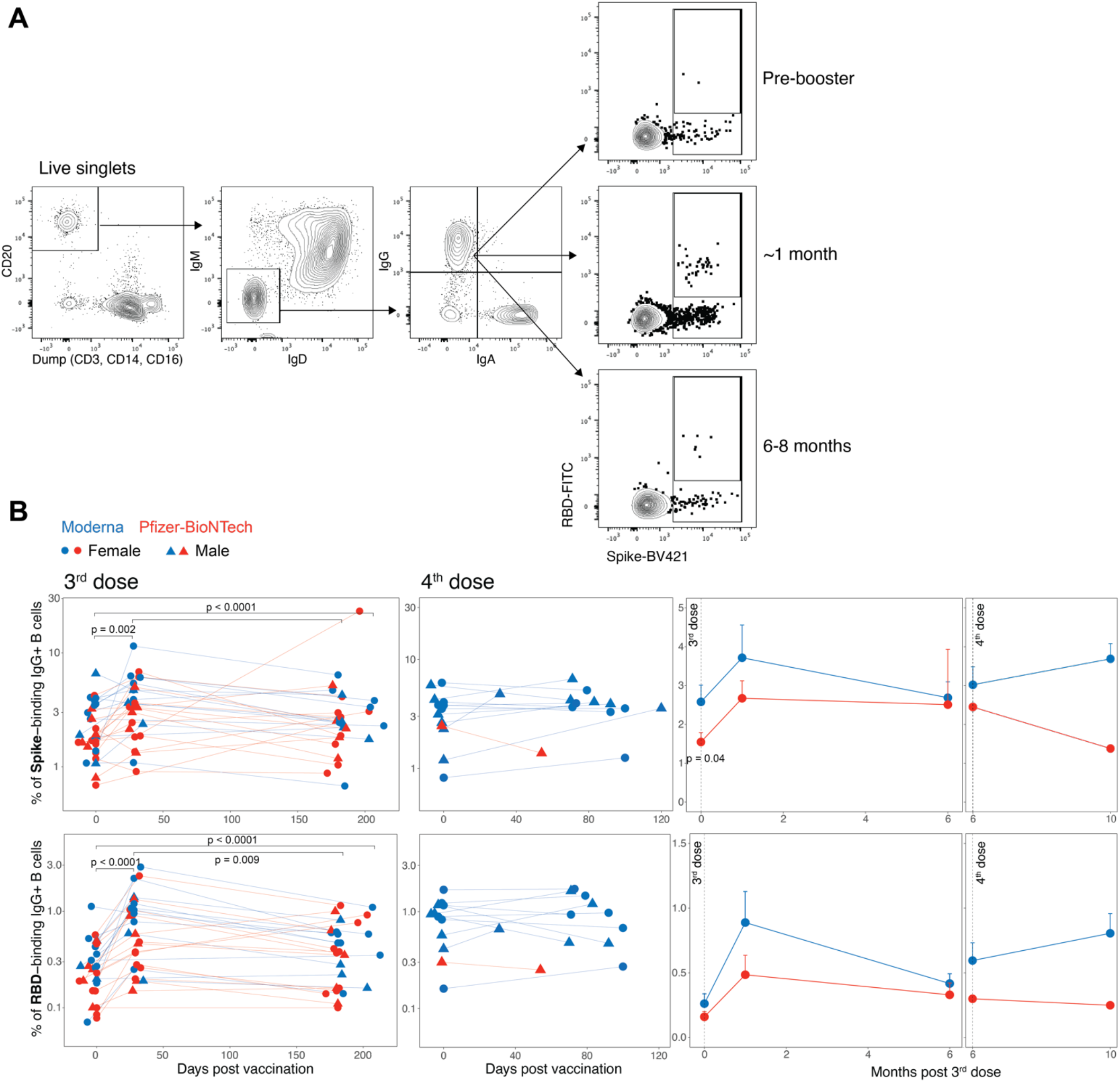
Memory B cell responses to the booster vaccination. **(A)** A representative flow cytometry profile to show the gating strategy to define Spike-specific B cell frequencies (Gated as live, CD20^+^ IgD^-^ IgM^-^ Spike^+^ RBD^+/-^ cells. (**B**) Frequency of Spike (top panel) or RBD (bottom panel) specific memory B cells relative to CD20^+^ IgD^-^ IgM^-^ B cells. Each symbol represents an individual (N = 28 after 3^rd^ dose and 13 after 4^th^ dose). The statistical differences between time points were determined using Wilcoxon matched-pairs signed rank test. The two graphs on the right show summary of the responses (Geomean + SEM). The statistical difference between the groups were determined using Mann-Whitney test.

We also measured T cell responses using intracellular cytokine staining (ICS) assay following a 6 h stimulation of peripheral blood mononuclear cells (PBMCs) with overlapping peptide pools spanning the Spike proteins of the ancestral and Omicron variants (Tarke et al., 2022). The booster vaccination induced significant CD4 T cell responses, primarily Th1-type **(Fig. 4A)** consistent with previous studies (Arunachalam *et al*., 2021; Sahin et al., 2020). The magnitude of response at ^7 days post-vaccination was significantly higher in response to mRNA1273 as compared to BNT162b2 vaccination. The response at the pre-booster time point was predominantly IL-2^+^ and TNF^+^ CD4 T cells with or without IFN-γ suggesting an establishment of potent memory T cells during previous vaccinations. While the frequency of cells producing IL-2 and TNF remained elevated after vaccination, the cells producing only IFN-γ were significantly induced by day 7 **(Fig. 4B)** suggesting potential differentiation of memory T cells to an effector phenotype. The magnitude of IFN-γ, TNF as well as IL-4 producing CD4 T cells, albeit at a much lower level, persisted until 6 months at a frequency higher than the pre-booster frequency **(Fig. 4B)**, but the proportion of cells producing multiple cytokines largely returned to the baseline state which comprised predominantly IL-2^+^ TNF^+^ +/- IFN-γ^+^ cells **(Fig. 4C)**. The CD4 T cells recognizing ancestral, or Omicron Spike antigens were relatively comparable suggesting conservation of T cell epitopes consistent with previous studies **(Fig. 4D)** (Gao et al., 2022; Keeton et al., 2022; Tarke *et al*., 2022). The booster vaccination also elicited CD8 T cell responses producing IFN-γ by day 7 that returned to pre-booster magnitude by 6 months **(Supplementary Fig. 3A, B)**. The T cell responses were also not significantly different between males and females **(Supplementary Fig. 3C, D)**. Collectively, these data demonstrate that the booster vaccination reactivates memory B and T cell responses and maintains the durable memory response elicited by prior vaccinations.

**Figure 4.**
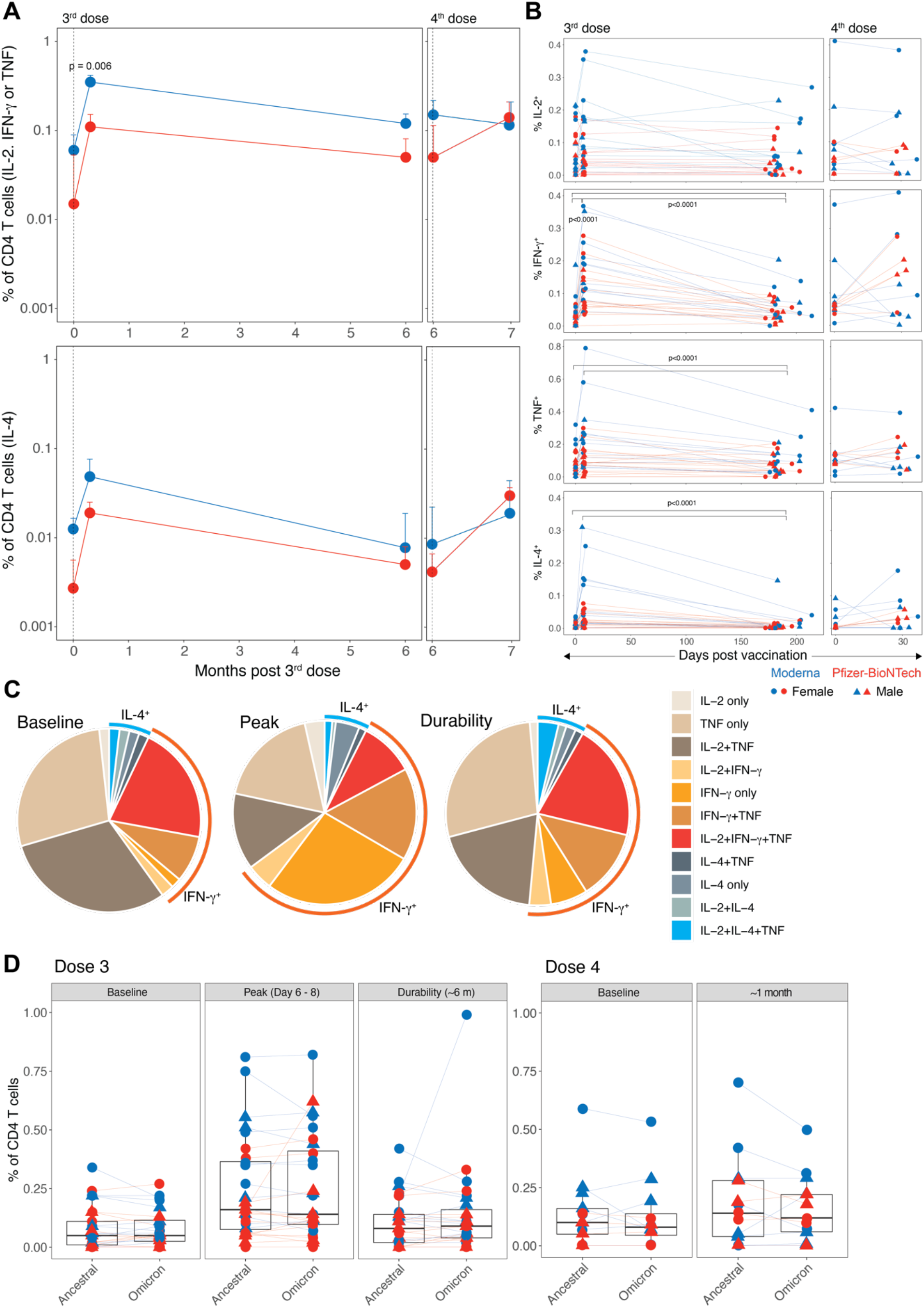
T cell responses induced by the booster mRNA vaccination. **(A)** A summary of the frequency of Spike-specific CD4 T cell responses secreting IL-2, IFN-γ, or TNF (Th1-type, top panel) and IL-4 (Th2-type, bottom panel). Median responses + SEM are plotted. The statistical difference between groups was determined using Mann-Whitney test. **(B)** Frequency of individuals cytokine producing Spike-specific CD4 T cells. Each symbol represents an individual (N = 28 after 3^rd^ dose and 13 after 4^th^ dose). The statistical significance between time points was calculated using Wilcoxon matched-pairs signed rank test. **(C)** Pie charts showing proportion of Spike-specific CD4 T cells producing one, two or three cytokines in response to the 3^rd^ dose vaccination. **(D)** Comparison of CD4 T cell frequencies (Th1-type producing IL-2, TNF or IFN-γ) between ancestral and Omicron viral strains measured at time points indicated on the plots.

## DISCUSSION

Messenger RNA vaccines stimulate durable immune memory (Goel *et al*., 2021); however, serum antibody responses steadily decline with a half-life of 56 – 66 days in the first 6 months after two doses (Doria-Rose *et al*., 2021; Pegu *et al*., 2021; Suthar *et al*., 2022) resulting in weak to undetectable nAb titers in many individuals. Concurrent with declining antibody titers, the effectiveness of vaccine-mediated protection against infection declines in the population (Feikin et al., 2022; Goldberg et al., 2021). The emergence of immune escape variants such as Omicron further reduces vaccine effectiveness mandating booster vaccinations (Hoffmann et al., 2022). Whether the durability of immune responses, in particular, the half-life of serum antibody titers improves after subsequent booster vaccinations, was the primary question addressed in this study.

Our data show that a booster vaccination (3^rd^ dose) increases the half-life of binding and neutralizing antibody titers only marginally when SARS-CoV-2 naïve participants were analyzed. While the increase in t_1/2_ of binding antibody titer was not statistically different, the increase in t_1/2_ of the nAb titer to 76 days from 56 days after the second vaccination in our previous study (Suthar *et al*., 2022) was statistically significant (p = 0.003 between the decay rates after the 2^nd^ and 3^rd^ vaccination, Wald test). The half-life improved further after the 4^th^ vaccination, although the analysis was only until 3 – 4 months and should be considered with caution. Whether the increase in half-life is truly an increase in durability associated with an increase in bone marrow plasma cells (BMPCs) warrants investigation. Evidence indicates the generation of BMPCs detectable at 6 months following two doses of BNT162b2; however, the magnitude was modest (Median 0.06% of total IgG-producing BMPCs) in contrast to 1.4% against seasonal influenza or 0.15% against Tetanus antigen (Kim et al., 2022). It is conceivable that BMPCs accumulate and increase in number after subsequent booster vaccinations, thus resulting in improving durability.

A secondary objective of our study was to determine the nAb response against emerging SARS-CoV-2 variants. The booster vaccination induced nAb response against Omicron subvariants BA.1, BA.5, and BA.2.75 consistent with previous reports (Edara *et al*., 2022; Qu et al., 2022a; Qu *et al*., 2022b; Shen et al., 2022). However, the responses against the most recently emerged variants BA.2.75.2 and BQ.1.1 with the concerning R346T mutation were strikingly diminished (35 – 49X). By 6 months, almost no one had a detectable titer against these variants. These findings, alongside our recent data indicating enhancement of breadth against these variants following a bivalent booster containing ancestral and BA.5 Spike (Davis-Gardner et al., 2022), suggest that monovalent boosters may be ineffective in protection against infection, and thus bivalent vaccines are required for subsequent boosters to improve the serological responses. In contrast, the memory T and B cell responses were durable after the booster vaccinations continuing to provide protection against severe disease.

The limitations of the study include a smaller cohort size as well as a shorter follow-up period (3 – 4 months as against 6 months) after the 4^th^ dose. It should also be noted that the individuals who received the 4^th^ dose were relatively older in comparison with the 3^rd^ dose vaccinees although all participants were immunocompetent. We also did not evaluate the effect of the bivalent booster in this study.

In summary, our comprehensive analysis of immune responses provides evidence that a lack of nAb responses against emerging variants despite the marginal improvement of durability underlies the declining efficacy of booster doses of mRNA vaccines and helps make decisions on subsequent booster vaccinations.

## Data Availability

All data produced in the present study are available upon reasonable request to the authors.

---

**Supplementary Fig. 1.**
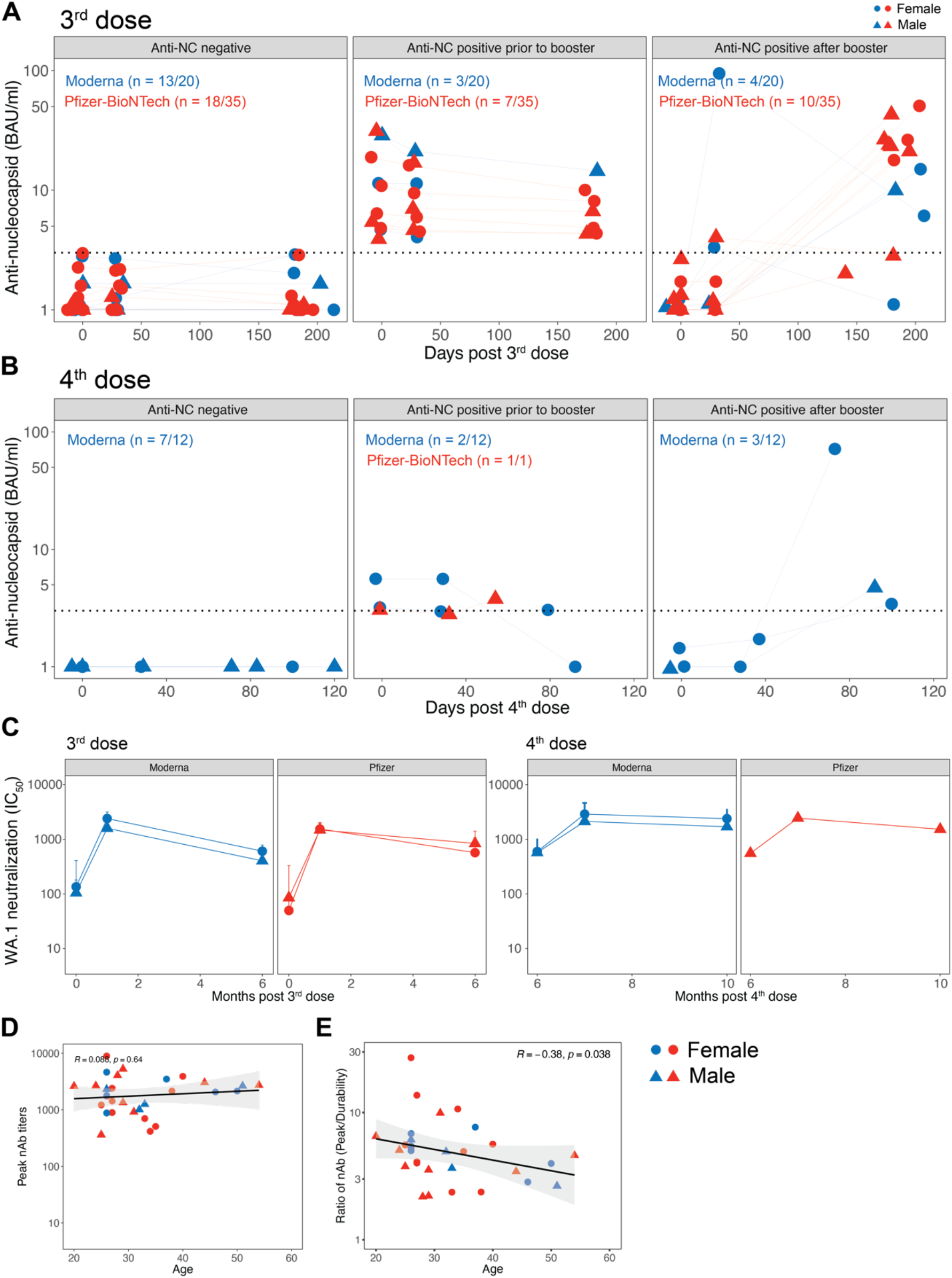
Serum antibody responses following booster mRNA vaccinations. **(A - B)** Anti-N antibody response measured in the sera of participants who received 3 **(A)** or 4 **(B)** doses. **(C)** A summary of neutralizing antibody response against the ancestral strain in males versus females. **(D - E)** Spearman’s correlation between age and neutralizing antibody titers against the ancestral strain at peak **(D)** or ratio of peak versus durability **(E)**.

**Supplementary Fig. 2.**
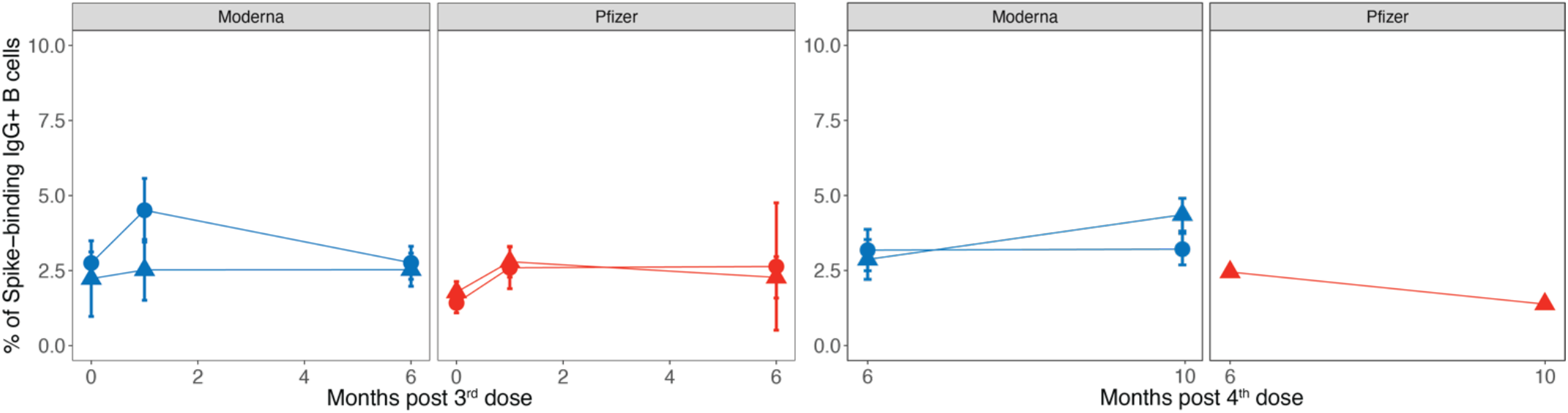
Memory B cell response following booster mRNA vaccinations. A summary of the frequency of Spike-binding memory B in males versus females.

**Supplementary Fig. 3.**
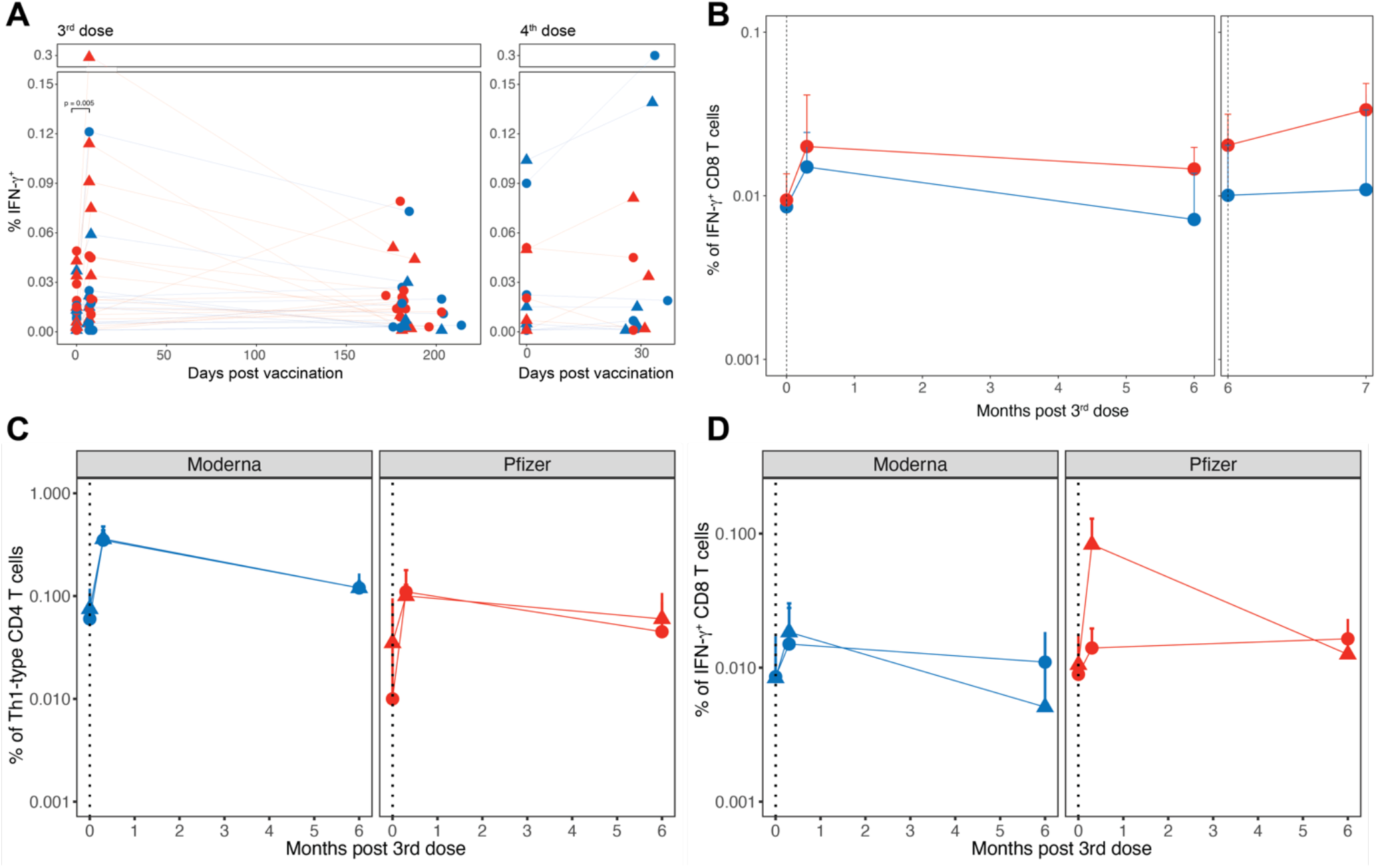
CD8 T cell response following booster mRNA vaccinations. **(A)** Frequency of Spike-specific CD8 T cells producing IFN-γ. **(B)** A summary of the frequency of Spike-specific CD8 T cells producing IFN-γ. Blue and red colors indicate Moderna and Pfizer-BioNTech vaccines. **(C – D)** A summary of the frequency of antigen-specific CD4 **(C)** and CD8 **(D)** T cells stratified by sex. Circles and triangles represent females and males, respectively.

**Supplementary Table 1.**

| **Participant** | **Age range** | **Gender** | **Race** | **Booster dose** | **Booster vaccine** | **COVID-19** | **Booster period** |
| --- | --- | --- | --- | --- | --- | --- | --- |
| 1 | 20-29 | Female | White | 3 | BNT162b2 | After | Sep - Dec 2021 |
| 2 | 40-49 | Female | White | 3 | BNT162b2 | No | Sep - Dec 2021 |
| 3 | 20-29 | Female | Black | 3 | BNT162b2 | No | Sep - Dec 2021 |
| 4 | 20-29 | Female | White | 3 | BNT162b2 | No | Sep - Dec 2021 |
| 5 | 40-49 | Male | Asian | 3 | BNT162b2 | No | Sep - Dec 2021 |
| 6 | 20-29 | Female | Asian | 3 | BNT162b2 | No | Sep - Dec 2021 |
| 7 | 20-29 | Male | Asian | 3 | BNT162b2 | After | Sep - Dec 2021 |
| 8 | 30-39 | Female | White | 3 | BNT162b2 | Before | Sep - Dec 2021 |
| 9 | 20-29 | Male | White | 3 | BNT162b2 | After | Sep - Dec 2021 |
| 10 | 20-29 | Male | White | 3 | BNT162b2 | Before | Sep - Dec 2021 |
| 11 | 30-39 | Female | White | 3 | BNT162b2 | No | Sep - Dec 2021 |
| 12 | 40-49 | Female | White | 3 | BNT162b2 | No | Sep - Dec 2021 |
| 13 | 20-29 | Male | White | 3 | BNT162b2 | After | Sep - Dec 2021 |
| 14 | 20-29 | Female | White | 3 | mRNA1273 | No | Sep - Dec 2021 |
| 15 | 20-29 | Male | Black | 3 | BNT162b2 | No | Sep - Dec 2021 |
| 16 | 20-29 | Female | White | 3 | BNT162b2 | No | Sep - Dec 2021 |
| 17 | 20-29 | Male | Asian | 3 | BNT162b2 | No | Sep - Dec 2021 |
| 18 | 20-29 | Female | Asian | 3 | BNT162b2 | Before | Sep - Dec 2021 |
| 19 | 30-39 | Male | White | 3 | BNT162b2 | No | Sep - Dec 2021 |
| 20 | 20-29 | Male | White | 3 | mRNA1273 | After | Sep - Dec 2021 |
| 21 | 20-29 | Male | White | 3 | BNT162b2 | After | Sep - Dec 2021 |
| 22 | 20-29 | Female | White | 3 | mRNA1273 | After | Sep - Dec 2021 |
| 23 | 20-29 | Male | Asian | 3 | BNT162b2 | No | Sep - Dec 2021 |
| 24 | 30-39 | Female | White | 3 | BNT162b2 | No | Sep - Dec 2021 |
| 25 | 30-39 | Female | White | 3 | BNT162b2 | No | Sep - Dec 2021 |
| 26 | 50-59 | Male | White | 3 | BNT162b2 | After | Sep - Dec 2021 |
| 27 | 60-69 | Female | White | 3 | BNT162b2 | After | Sep - Dec 2021 |
| 28 | 50-59 | Female | White | 3 | BNT162b2 | After | Sep - Dec 2021 |
| 29 | 40-49 | Male | White | 3 | BNT162b2 | After | Sep - Dec 2021 |
| 30 | 30-39 | Female | White | 3 | BNT162b2 | No | Sep - Dec 2021 |
| 31 | 50-59 | Male | White | 3 | mRNA1273 | Before | Sep - Dec 2021 |
| 32 | 20-29 | Male | Asian | 3 | BNT162b2 | No | Sep - Dec 2021 |
| 33 | 30-39 | Male | White | 3 | mRNA1273 | No | Sep - Dec 2021 |
| 34 | 20-29 | Female | Mixed | 3 | mRNA1273 | No | Sep - Dec 2021 |
| 35 | 50-59 | Female | Black | 3 | mRNA1273 | No | Sep - Dec 2021 |
| 36 | 70-79 | Male | White | 3 | mRNA1273 | No | Sep - Dec 2021 |
| 37 | 20-29 | Male | White | 3 | BNT162b2 | After | Sep - Dec 2021 |
| 38 | 30-39 | Female | White | 3 | BNT162b2 | No | Sep - Dec 2021 |
| 39 | 60-69 | Female | White | 3 | BNT162b2 | No | Sep - Dec 2021 |
| 40 | 20-29 | Male | White | 3 | BNT162b2 | Before | Sep - Dec 2021 |
| 41 | 30-39 | Female | White | 3 | BNT162b2 | Before | Sep - Dec 2021 |
| 42 | 30-39 | Female | White | 3 | mRNA1273 | After | Sep - Dec 2021 |
| 43 | 30-39 | Female | White | 3 | mRNA1273 | No | Sep - Dec 2021 |
| 44 | 50-59 | Female | White | 3 | mRNA1273 | No | Sep - Dec 2021 |
| 45 | 20-29 | Female | White | 3 | BNT162b2 | Before | Sep - Dec 2021 |
| 46 | 20-29 | Female | White | 3 | mRNA1273 | No | Sep - Dec 2021 |
| 47 | 20-29 | Female | Asian | 3 | mRNA1273 | No | Sep - Dec 2021 |
| 48 | 70-79 | Male | Black | 3 | mRNA1273 | No | Sep - Dec 2021 |
| 49 | 60-69 | Female | Black | 3 | mRNA1273 | Before | Sep - Dec 2021 |
| 50 | 60-69 | Female | White | 3 | mRNA1273 | No | Sep - Dec 2021 |
| 51 | 30-39 | Male | White | 3 | mRNA1273 | No | Sep - Dec 2021 |
| 52 | 40-49 | Female | White | 3 | mRNA1273 | After | Sep - Dec 2021 |
| 53 | 30-39 | Female | White | 3 | mRNA1273 | No | Jan - Apr 2022 |
| 54 | 30-39 | Female | Black | 3 | mRNA1273 | Before | Jan - Apr 2022 |
| 55 | 30-39 | Male | White | 3 | BNT162b2 | Before | Jan - Apr 2022 |
| 56 | 70-79 | Male | White | 4 | mRNA1273 | No | Jan - Apr 2022 |
| 57 | 50-59 | Male | White | 4 | mRNA1273 | No | Jan - Apr 2022 |
| 58 | 60-69 | Female | Black | 4 | mRNA1273 | No | Jan - Apr 2022 |
| 59 | 60-69 | Female | Black | 4 | mRNA1273 | After | Jan - Apr 2022 |
| 60 | 70-79 | Female | White | 4 | mRNA1273 | Before | May - Aug 2022 |
| 61 | 60-69 | Male | White | 4 | mRNA1273 | After | May - Aug 2022 |
| 62 | 60-69 | Male | White | 4 | mRNA1273 | No | May - Aug 2022 |
| 63 | 70-79 | Male | White | 4 | mRNA1273 | No | May - Aug 2022 |
| 64 | 60-69 | Male | Black | 4 | mRNA1273 | No | May - Aug 2022 |
| 65 | 80-89 | Female | White | 4 | mRNA1273 | Before | May - Aug 2022 |
| 66 | 60-69 | Female | White | 4 | mRNA1273 | No | May - Aug 2022 |
| 67 | 70-79 | Female | White | 4 | mRNA1273 | After | May - Aug 2022 |
| 68 | 60-69 | Male | White | 4 | BNT162b2 | Before | May - Aug 2022 |

## MATERIALS AND METHODS

### Study participants

All the participants of the study were recruited at the Hope Clinic of the Emory Vaccine Center. The participant details are provided in Supplementary Table 1 with deidentified information. The study was reviewed and approved by Emory IRB (Study 00002061).

### Electrochemiluminescence (ECL) binding enzyme-linked immunosorbent assay for anti-Spike and anti-nucleocapsid antibodies

Anti-Spike and anti-nucleocapsid protein IgG titers were measured using V-plex SARS-CoV-2 panels 29 (Cat #K15624U) and 2 (Cat #K15383U), respectively, from Mesoscale. The assay was performed as per the manufacturer’s instructions. Briefly, the multi-spot 96 well plates were blocked in 0.15 ml of blocking solution with shaking at 700 rpm at room temperature. After 30 minutes of incubation, 50 μl of serum samples were diluted in antibody diluent solution, and serially diluted calibrator solution were added to each plate in the designated wells and incubated at room temperature for 2 hours with shaking. After 2 hours of incubation, the plates were washed and 50 μl of Sulfo-tag conjugated anti-IgG was added, and the plates were incubated at room temperature for 1 hour. After incubation, the plates were washed, and 0.15 ml of MSD-Gold read buffer was added. The plates were immediately read using the MSD instrument. The unknown concentrations were extrapolated using a standard curve drawn using the calibrators in each plate and presented as relative MSD Arbitrary Units AU/ml or BAU/ml.

### Viruses and cells for Focus Reduction Neutralization Test (FRNT)

VeroE6-TMPRSS2 cells were described previously and cultured in complete DMEM (DMEM with 10% FBS + Penicillin/Streptomycin) in the presence of Gibco Puromycin 10 mg/ml (#A11138-03). nCoV/USA_WA1/2020 (WA/1) was propagated from an infectious SARS-CoV-2 clone as previously described (Xie et al., 2020). icSARS-CoV-2 was passaged once to generate a working stock. The BA.1 isolate has been previously described (Edara Cell Reports 2022). Omicron subvariants were isolated from residual nasal swabs: BA.5 isolate (EPI_ISL_13512579), provided by Dr. Richard Webby (St Jude Children’s Research Hospital), BA.2.75.2 (EPI_ISL_15146622), BQ.1.1 isolate (EPI_ISL_15196219), and BA.2.75 isolate (EPI_ISL_14393635) provided by Dr. Benjamin Pinsky (Stanford University). All variants were plaque purified and propagated once in VeroE6-TMPRSS2 cells to generate working stocks. Viruses were deep-sequenced and confirmed as previously described (Edara et al., 2021).

FRNT assays were performed as previously described (Edara *et al*., 2022; Edara *et al*., 2021; Vanderheiden et al., 2020). Briefly, samples were diluted three-fold in eight serial dilutions using DMEM in duplicates with an initial dilution of 1:10 in a total volume of 60 μl. Serially diluted samples were incubated with an equal volume of SARS-CoV-2 (100 to 200 foci per well based on the target cell) at 37ºC for 45 minutes in a round-bottomed 96-well culture plate. The antibody-virus mixture was then added to VeroE6-TMPRSS2 cells and incubated at 37ºC for 1 hour. Post-incubation, the antibody-virus mixture was removed and 100 μl of pre-warmed 0.85% methylcellulose (Sigma-Aldrich, #M0512-250G) overlay was added to each well. Plates were incubated at 37ºC for either 18 to 40 hours and the methylcellulose overlay was removed and washed six times with PBS. Cells were fixed with 2% paraformaldehyde in PBS for 30 minutes. Following fixation, plates were washed twice with PBS and permeabilization buffer (0.1% Bovine serum albumin (BSA) and 0.1% Saponin in PBS) was added to permeabilized cells for at least 20 minutes. Cells were incubated with an anti-SARS-CoV Spike protein primary antibody directly conjugated to Alexa fluor (AF)-647 (CR3022-AF647) for 4 hours at room temperature or overnight at 4ºC. Cells were washed three times in PBS and foci were visualized on an ELISPOT reader. Antibody neutralization was quantified by counting the number of foci for each sample using the Viridot program (Katzelnick et al., 2018). The neutralization titers were calculated as follows: 1 - (ratio of the mean number of foci in the presence of serum and foci at the highest dilution of respective serum sample). Each specimen was tested in duplicate. The FRNT-50 titers were interpolated using a 4-parameter nonlinear regression in GraphPad Prism 9.2.0. Samples that do not neutralize at the limit of detection at 50% are plotted at 10 and was used for geometric mean and fold-change calculations.

### Antibody half-life calculations

Mixed-effects models implemented in MonolixSuite 2021R1 (Lixoft) were used to estimate the corresponding half-lives of antigen-specific antibodies. The exponential decay model dAb/dt=-*k*\*Ab was fitted to the longitudinal data starting from day 21 after third of fourth vaccine doses, where Ab is the antibody concentration and *k* is the exponential decay. The corresponding half-lives were calculated as t_1/2_=ln(2)/*k*. The individual-level parameters were lognormally distributed for the initial antibody concentration (^1 month) and normally distributed for the decay rate *k* with an assumption of no correlations between the random effects. We assumed multiplicative independent lognormal observation error. The estimation of the population parameters was performed using the Stochastic Approximation Expectation-Maximization (SAEM) algorithm.

### Spike protein-specific memory B cell staining

Cryopreserved PBMCs were thawed and washed twice with 10 ml of FACS buffer (1 x PBS containing 2% FBS and 1 mM EDTA) and resuspended in 100 μl of PBS containing Zombie UV live/dead dye at 1:200 dilution (BioLegend #423108) and incubate at room temperature for 15 minutes. Following washing, cells were incubated with an antibody cocktail for 1 hour protected from light on ice. The following antibodies were used: IgD phycoerythrin (PE, Southern Biotech #2030-09), IgM peridinin chlorophyll protein (PerCP)-Cy5.5 (BioLegend #314512), CD20 allophycocyanin (APC)-H7 (BD Biosciences #560734), CD27 PE-Cy7 (BioLegend #302838), CD14 brilliant violet (BV) 650 (BioLegend #301836), CD16 BV650 (BioLegend #302042), IgG brilliant ultraviolet (BUV) 496 (BD Biosciences #741172), CD3 BV650 (BD Biosciences #563916), CD21 PE-CF594 (BD Biosciences #563474), Alexa Fluor 647 labeled Omicron Spike protein (SinoBiological #40589-V08H26), BV605-labelled Omicron RBD protein (SinoBiological), BV421 labeled ancestral Spike protein (SinoBiological #40589-V27B-B) and FITC-labelled ancestral RBD protein (SinoBiological). All antibodies were used as the manufacturer’s instruction and the final concentration of each probe was 0.1 μg/ml. Cells were washed twice in FACS buffer and immediately acquired on a BD FACS Aria III. Flowjo software v10 (TreeStar Inc.) was used for data analysis.

### Intracellular cytokine staining assay

Antigen-specific T cell responses were measured using the intracellular cytokine staining assay. Live frozen PBMCs were revived, counted, and resuspended at a density of 2 × 10^6^ live cells per ml in complete Rosewell Park Memorial Institute (RPMI)-1640 (RPMI-1640 supplemented with 10% FBS and Penicillin-streptomycin) and rested for 6 h at 37°C in a CO_2_ incubator. After the incubation, the cells were washed once and resuspended at a density of 12 - 15 × 10^6^ per ml in complete RPMI-1640 and 100 μl of cell suspension containing 1.2 – 1.5 × 10^6^ cells was added to each well of a 96-well round-bottomed tissue culture plate. Each sample was treated with 2 or 3 conditions depending on cell numbers: no stimulation or a peptide pool spanning the Spike protein of the ancestral Wu strain or Omicron BA.1 variant (where cell numbers permitted) in the presence of 1 μg ml^−1^ of anti-CD28 (clone CD28.2, BD Biosciences) and anti-CD49d (clone 9F10, BD Biosciences) as well as anti-CXCR3 and anti-CXCR5. The details of peptide synthesis and purity are described previously (Tarke *et al*., 2022). Briefly, the peptide pools were 15-mer peptides with 10-mer overlaps spanning the entire Spike protein sequence of each variant. The amino acids in the variant peptide pools that vary from the ancestral Spike protein sequence are provided in Table S3 of Tarke *et al*. (Tarke *et al*., 2022). Each peptide was dissolved at a concentration of 20 mg/ml in dimethyl sulfoxide (DMSO) and individual peptides were pooled to prepare each variant-specific peptide pool following sequential lyophilization, as previously reported (Tarke *et al*., 2022). Each peptide pool contained 253 peptides and was resuspended in DMSO at a concentration of 1 mg/ml. PBMCs were stimulated at a final concentration of 1 μg/ml of each peptide in the final reaction with an equimolar amount of DMSO (0.5% v/v in 0.2 ml total reaction volume) as negative control. The samples were incubated at 37°C in CO2 incubators for 2 hours before addition of 10 μg ml^−1^ brefeldin A. The cells were incubated for an additional 4 hours. The cells were washed with PBS and stained with Zombie UV fixable viability dye (BioLegend). The cells were washed with PBS containing 5% FBS, before the addition of surface antibody cocktail. The cells were stained for 20 minutes at 4°C in 100 μl volume. Subsequently, the cells were washed, fixed and permeabilized with cytofix/cytoperm buffer (BD Biosciences #555028) for 20 minutes. The permeabilized cells were stained with intracellular cytokine staining antibodies for 20 minutes at room temperature in 1× perm/wash buffer (BD Biosciences #555028). The details of antibody panel used in the assay were described previously in (Arunachalam et al., 2022). Cells were then washed twice with perm/wash buffer and once with staining buffer before acquisition using the BD Symphony Flow Cytometer and the associated BD FACS Diva software. All flow cytometry data were analysed using Flowjo software v10 (TreeStar Inc.).

### Statistical analysis

The difference between any two groups at a time point was measured using a two-tailed nonparametric Mann–Whitney unpaired rank-sum test. The difference between time points within a group was measured using a Wilcoxon matched-pairs signed-rank test. The correlations were Spearman’s correlations based on ranks. All statistical analyses were performed using GraphPad Prism v.9.0.0 or R version 3.6.1. We compared exponential decay rates for binding and neutralizing antibodies after 2^nd^ and 3^rd^ vaccine doses using Wald test implemented in R. For binding antibodies, exponential decay rates estimated using mixed-effects models after 2^nd^ dose (Suthar *et al*., 2022) and after 3^rd^ dose with breakthrough cases removed were 0.0124 (se=0.000644) per day and 0.00913 (se=0.000892) per day, respectively. For neutralizing antibody titers, exponential decay rates after 2^nd^ dose (Suthar *et al*., 2022) and after 3^rd^ dose with breakthrough cases removed were 0.0124 (se=0.000644) per day and 0.00913 (se=0.000892) per day, respectively. All the figures were made in GraphPad Prism or R and organized in Adobe Illustrator.

## Acknowledgments

This study was supported by the NIH grants U19 AI057266 and U19 AI167903, Bill and Melinda Gates Foundation, Open Philanthropy, the Violetta L. Horton and Soffer Endowments, and contributions from an anonymous donor, to B.P. This work was supported in part by grants (NIH P51OD011132, 1U54CA260563, HHSN272201400004C, NIH/NIAID CEIRR under contract 75N93021C00017 to Emory University) from the National Institute of Allergy and Infectious Diseases (NIAID), National Institutes of Health (NIH), Woodruff Health Sciences Center 2020 COVID-19 CURE Award to M.S.S. Additional NIH support was provided under Contract No. 75N93021C00016 to A.G. and A.G. and U1 AI141995 and U19 AI118626 to A.S. We thank all the members of the Hope Clinic and the study participants for the valuable samples. The schematic was made using BioRender.

## Author contributions

B.P. conceived the study; P.S.A. and B.P. designed the study and were responsible for the overall conduct of the study; H.S., C.H., K.B., S.B., M.L., M. Litcack., C.L., and N.R. collected clinical samples; P.S.A., and Y.F. performed binding antibody assays; L.L., B.W., and M.E. performed neutralization assays; P.S.A., Y.F., M.H., and H.S.H. performed memory T and B cell assays; V.I.Z. calculated antibody half-lives; A.G. and A.S. provided peptide pools for T cell assays; N.R., M.S.S., and B.P. supervised the experiments; P.S.A. formally analyzed all the datasets and prepared the figures; P.S.A., and B.P. wrote the manuscript with suggestions and assistance from all co-authors. All the authors read and accepted the final contents of the manuscript.

## References

Ailsworth, S.M., Keshavarz, B., Richards, N.E., Workman, L.J., Murphy, D.D., Nelson, M.R., Platts-Mills, T.A.E., and Wilson, J.M. (2022). Enhanced SARS-CoV-2 IgG durability following COVID-19 mRNA booster vaccination and comparison of BNT162b2 with mRNA-1273. Ann Allergy Asthma Immunol. 10.1016/j.anai.2022.10.003.

Andrews, N., Stowe, J., Kirsebom, F., Toffa, S., Rickeard, T., Gallagher, E., Gower, C., Kall, M., Groves, N., O’Connell, A.M., et al. (2022). Covid-19 Vaccine Effectiveness against the Omicron (B.1.1.529) Variant. N Engl J Med. 10.1056/NEJMoa2119451.

Arunachalam, P.S., Feng, Y., Ashraf, U., Hu, M., Walls, A.C., Edara, V.V., Zarnitsyna, V.I., Aye, P.P., Golden, N., Miranda, M.C., et al. (2022). Durable protection against the SARS-CoV-2 Omicron variant is induced by an adjuvanted subunit vaccine. Sci Transl Med 14, eabq4130. 10.1126/scitranslmed.abq4130.

Arunachalam, P.S., Scott, M.K.D., Hagan, T., Li, C., Feng, Y., Wimmers, F., Grigoryan, L., Trisal, M., Edara, V.V., Lai, L., et al. (2021). Systems vaccinology of the BNT162b2 mRNA vaccine in humans. Nature. 10.1038/s41586-021-03791-x.

Collie, S., Nayager, J., Bamford, L., Bekker, L.G., Zylstra, M., and Gray, G. (2022). Effectiveness and Durability of the BNT162b2 Vaccine against Omicron Sublineages in South Africa. N Engl J Med 387, 1332–1333. 10.1056/NEJMc2210093.

Davis-Gardner, M.E., Lai, L., Wali, B., Samaha, H., Solis, D., Lee, M., Porter-Morrison, A., Hentenaar, I.T., Yamamoto, F., Godbole, S., et al. (2022). mRNA bivalent booster enhances neutralization against BA.2.75.2 and BQ.1.1. bioRxiv. 10.1101/2022.10.31.514636.

Doria-Rose, N., Suthar, M.S., Makowski, M., O’Connell, S., McDermott, A.B., Flach, B., Ledgerwood, J.E., Mascola, J.R., Graham, B.S., Lin, B.C., et al. (2021). Antibody Persistence through 6 Months after the Second Dose of mRNA-1273 Vaccine for Covid-19. N Engl J Med. 10.1056/NEJMc2103916.

Edara, V.V., Manning, K.E., Ellis, M., Lai, L., Moore, K.M., Foster, S.L., Floyd, K., Davis-Gardner, M.E., Mantus, G., Nyhoff, L.E., et al. (2022). mRNA-1273 and BNT162b2 mRNA vaccines have reduced neutralizing activity against the SARS-CoV-2 omicron variant. Cell Rep Med 3, 100529. 10.1016/j.xcrm.2022.100529.

Edara, V.V., Pinsky, B.A., Suthar, M.S., Lai, L., Davis-Gardner, M.E., Floyd, K., Flowers, M.W., Wrammert, J., Hussaini, L., Ciric, C.R., et al. (2021). Infection and Vaccine-Induced Neutralizing-Antibody Responses to the SARS-CoV-2 B.1.617 Variants. N Engl J Med 385, 664–666. 10.1056/NEJMc2107799.

Feikin, D.R., Higdon, M.M., Abu-Raddad, L.J., Andrews, N., Araos, R., Goldberg, Y., Groome, M.J., Huppert, A., O’Brien, K.L., Smith, P.G., et al. (2022). Duration of effectiveness of vaccines against SARS-CoV-2 infection and COVID-19 disease: results of a systematic review and meta-regression. Lancet 399, 924–944. 10.1016/S0140-6736(22)00152-0.

Gao, Y., Cai, C., Grifoni, A., Muller, T.R., Niessl, J., Olofsson, A., Humbert, M., Hansson, L., Osterborg, A., Bergman, P., et al. (2022). Ancestral SARS-CoV-2-specific T cells cross-recognize the Omicron variant. Nat Med. 10.1038/s41591-022-01700-x.

Garcia-Beltran, W.F., St Denis, K.J., Hoelzemer, A., Lam, E.C., Nitido, A.D., Sheehan, M.L., Berrios, C., Ofoman, O., Chang, C.C., Hauser, B.M., et al. (2022). mRNA-based COVID-19 vaccine boosters induce neutralizing immunity against SARS-CoV-2 Omicron variant. Cell 185, 457–466 e454. 10.1016/j.cell.2021.12.033.

Gilboa, M., Regev-Yochay, G., Mandelboim, M., Indenbaum, V., Asraf, K., Fluss, R., Amit, S., Mendelson, E., Doolman, R., Afek, A., et al. (2022). Durability of Immune Response After COVID-19 Booster Vaccination and Association With COVID-19 Omicron Infection. JAMA Netw Open 5, e2231778. 10.1001/jamanetworkopen.2022.31778.

Goel, R.R., Painter, M.M., Apostolidis, S.A., Mathew, D., Meng, W., Rosenfeld, A.M., Lundgreen, K.A., Reynaldi, A., Khoury, D.S., Pattekar, A., et al. (2021). mRNA Vaccination Induces Durable Immune Memory to SARS-CoV-2 with Continued Evolution to Variants of Concern. bioRxiv. 10.1101/2021.08.23.457229.

Goldberg, Y., Mandel, M., Bar-On, Y.M., Bodenheimer, O., Freedman, L., Haas, E.J., Milo, R., Alroy-Preis, S., Ash, N., and Huppert, A. (2021). Waning Immunity after the BNT162b2 Vaccine in Israel. N Engl J Med. 10.1056/NEJMoa2114228.

Hoffmann, M., Kruger, N., Schulz, S., Cossmann, A., Rocha, C., Kempf, A., Nehlmeier, I., Graichen, L., Moldenhauer, A.S., Winkler, M.S., et al. (2022). The Omicron variant is highly resistant against antibody-mediated neutralization: Implications for control of the COVID-19 pandemic. Cell 185, 447–456 e411. 10.1016/j.cell.2021.12.032.

Katzelnick, L.C., Coello Escoto, A., McElvany, B.D., Chavez, C., Salje, H., Luo, W., Rodriguez-Barraquer, I., Jarman, R., Durbin, A.P., Diehl, S.A., et al. (2018). Viridot: An automated virus plaque (immunofocus) counter for the measurement of serological neutralizing responses with application to dengue virus. PLoS Negl Trop Dis 12, e0006862. 10.1371/journal.pntd.0006862.

Keeton, R., Tincho, M.B., Ngomti, A., Baguma, R., Benede, N., Suzuki, A., Khan, K., Cele, S., Bernstein, M., Karim, F., et al. (2022). T cell responses to SARS-CoV-2 spike cross-recognize Omicron. Nature. 10.1038/s41586-022-04460-3.

Kim, W., Zhou, J.Q., Horvath, S.C., Schmitz, A.J., Sturtz, A.J., Lei, T., Liu, Z., Kalaidina, E., Thapa, M., Alsoussi, W.B., et al. (2022). Germinal centre-driven maturation of B cell response to mRNA vaccination. Nature 604, 141–145. 10.1038/s41586-022-04527-1.

Lustig, Y., Gonen, T., Meltzer, L., Gilboa, M., Indenbaum, V., Cohen, C., Amit, S., Jaber, H., Doolman, R., Asraf, K., et al. (2022). Superior immunogenicity and effectiveness of the third compared to the second BNT162b2 vaccine dose. Nat Immunol 23, 940–946. 10.1038/s41590-022-01212-3.

Moreira, E.D., Jr., Kitchin, N., Xu, X., Dychter, S.S., Lockhart, S., Gurtman, A., Perez, J.L., Zerbini, C., Dever, M.E., Jennings, T.W., et al. (2022). Safety and Efficacy of a Third Dose of BNT162b2 Covid-19 Vaccine. N Engl J Med 386, 1910–1921. 10.1056/NEJMoa2200674.

Pajon, R., Doria-Rose, N.A., Shen, X., Schmidt, S.D., O’Dell, S., McDanal, C., Feng, W., Tong, J., Eaton, A., Maglinao, M., et al. (2022). SARS-CoV-2 Omicron Variant Neutralization after mRNA-1273 Booster Vaccination. N Engl J Med. 10.1056/NEJMc2119912.

Pegu, A., O’Connell, S., Schmidt, S.D., O’Dell, S., Talana, C.A., Lai, L., Albert, J., Anderson, E., Bennett, H., Corbett, K.S., et al. (2021). Durability of mRNA-1273 vaccine-induced antibodies against SARS-CoV-2 variants. Science. 10.1126/science.abj4176.

Qu, P., Evans, J.P., Faraone, J., Zheng, Y.M., Carlin, C., Anghelina, M., Stevens, P., Fernandez, S., Jones, D., Lozanski, G., et al. (2022a). Distinct Neutralizing Antibody Escape of SARS-CoV-2 Omicron Subvariants BQ.1, BQ.1.1, BA.4.6, BF.7 and BA.2.75.2. bioRxiv. 10.1101/2022.10.19.512891.

Qu, P., Faraone, J.N., Evans, J.P., Zheng, Y.M., Yu, L., Ma, Q., Carlin, C., Lozanski, G., Saif, L.J., Oltz, E.M., et al. (2022b). Durability of Booster mRNA Vaccine against SARS-CoV-2 BA.2.12.1, BA.4, and BA.5 Subvariants. N Engl J Med 387, 1329–1331. 10.1056/NEJMc2210546.

Regev-Yochay, G., Gonen, T., Gilboa, M., Mandelboim, M., Indenbaum, V., Amit, S., Meltzer, L., Asraf, K., Cohen, C., Fluss, R., et al. (2022). Efficacy of a Fourth Dose of Covid-19 mRNA Vaccine against Omicron. N Engl J Med 386, 1377–1380. 10.1056/NEJMc2202542.

Sahin, U., Muik, A., Derhovanessian, E., Vogler, I., Kranz, L.M., Vormehr, M., Baum, A., Pascal, K., Quandt, J., Maurus, D., et al. (2020). COVID-19 vaccine BNT162b1 elicits human antibody and TH1 T-cell responses. Nature. 10.1038/s41586-020-2814-7.

Shen, X., Chalkias, S., Feng, J., Chen, X., Zhou, H., Marshall, J.C., Girard, B., Tomassini, J.E., Aunins, A., Das, R., and Montefiori, D.C. (2022). Neutralization of SARS-CoV-2 Omicron BA.2.75 after mRNA-1273 Vaccination. N Engl J Med 387, 1234–1236. 10.1056/NEJMc2210648.

Sievers, B.L., Chakraborty, S., Xue, Y., Gelbart, T., Gonzalez, J.C., Cassidy, A.G., Golan, Y., Prahl, M., Gaw, S.L., Arunachalam, P.S., et al. (2022). Antibodies elicited by SARS-CoV-2 infection or mRNA vaccines have reduced neutralizing activity against Beta and Omicron pseudoviruses. Sci Transl Med 14, eabn7842. 10.1126/scitranslmed.abn7842.

Suthar, M., Arunachalam, P., Hu, M., Reis, N., Trisal, M., Raeber, O., Chinthrajah, S., Davis-Gardner, M., Manning, K., Mudvari, P., et al. (2022). Durability of immune responses to the BNT162b2 mRNA vaccine. Med 3, 25–27. https://doi.org/10.1016/j.medj.2021.12.005.

Tarke, A., Coelho, C.H., Zhang, Z., Dan, J.M., Yu, E.D., Methot, N., Bloom, N.I., Goodwin, B., Phillips, E., Mallal, S., et al. (2022). SARS-CoV-2 vaccination induces immunological T cell memory able to cross-recognize variants from Alpha to Omicron. Cell 185, 847–859 e811. 10.1016/j.cell.2022.01.015.

Vanderheiden, A., Edara, V.V., Floyd, K., Kauffman, R.C., Mantus, G., Anderson, E., Rouphael, N., Edupuganti, S., Shi, P.Y., Menachery, V.D., et al. (2020). Development of a Rapid Focus Reduction Neutralization Test Assay for Measuring SARS-CoV-2 Neutralizing Antibodies. Curr Protoc Immunol 131, e116. 10.1002/cpim.116.

Xie, X., Muruato, A., Lokugamage, K.G., Narayanan, K., Zhang, X., Zou, J., Liu, J., Schindewolf, C., Bopp, N.E., Aguilar, P.V., et al. (2020). An Infectious cDNA Clone of SARS-CoV-2. Cell Host Microbe 27, 841–848 e843. 10.1016/j.chom.2020.04.004.

